# Matrix matters: head-to-head concordance of serum and plasma for NULISAseq CNS Disease Panel

**DOI:** 10.64898/2026.06.21.26356186

**Authors:** Tommaso Merati, Chiara Tolassi, Alessandro Rondina, Irene Girotto, Marianna Bertoni, Eileen Mac Sweeney, Andrea Toja, Eqrem Rusi, Caterina Martinuzzo, Andrea Pilotto, Alessandro Padovani

**Author notes:** Tommaso Merati and Chiara Tolassi contributed equally to this work and share first authorship. Corresponding author: Tommaso Merati, Neurology Unit, Department of Clinical and Experimental Sciences, University of Brescia and ASST Spedali Civili, Piazzale Spedali Civili 1, 25123 Brescia, Italy.

## Abstract

Blood-based proteomic profiling is now widely applied in neurodegenerative and neuroinflammatory disease, yet the choice between serum and plasma remains poorly characterised for high-multiplex platforms. Many legacy biobanks hold mainly serum, whereas most current NUcleic-acid-Linked Immuno-Sandwich Assay (NULISA) studies use plasma. We compared the 130-protein NULISAseq central nervous system (CNS) Disease Panel head-to-head in matched serum and plasma collected at the same draw from 62 participants (30 neurodegenerative, 19 demyelinating, 13 healthy controls). Agreement was measured with Spearman correlation (rho), Lin’s concordance correlation coefficient (CCC), the intraclass correlation coefficient (ICC) and the mean paired serum-to-plasma difference (dNPQ). Concordance was moderate to high: 123 of 130 proteins reached significance and 18 reached rho >= 0.90, with a median rho of 0.72 (range 0.10-0.988). Proteins fell into three tiers. Cytoskeletal markers (NEFH rho=0.988; NEFL rho=0.947) and glial GFAP (rho=0.949, |dNPQ|<0.5) were interchangeable between matrices. Phosphorylated tau (pTau) species retained excellent rank concordance but carried a systematic plasma-greater-than-serum offset (pTau-181 rho=0.869, dNPQ=+0.67; pTau-217 rho=0.846, dNPQ=+0.64; pTau-231 rho=0.885, dNPQ=+0.89). Platelet-derived analytes (CD40LG rho=0.102, dNPQ=-4.74; BDNF rho=0.223, dNPQ=-2.69) and intracellular synaptic proteins (NRGN, SNAP25, ENO2) diverged markedly. For most clinically relevant neurodegeneration markers, especially cytoskeletal and glial proteins, serum is a valid substitute for plasma; absolute thresholds for phosphorylated tau and amyloid peptides require matrix-specific calibration, and platelet-sensitive analytes cannot be compared across matrices without strictly standardised pre-analytical conditions.

**Main Points:**

- Across the 130-protein NULISAseq CNS Disease Panel, serum and plasma showed moderate-to-high overall concordance (median Spearman rho 0.72; 123/130 proteins significant).
- Neurofilaments (NEFH, NEFL) and GFAP were interchangeable between matrices, whereas phosphorylated tau species retained rank order but required matrix-specific calibration of absolute levels.
- Platelet-derived (CD40LG, BDNF) and intracellular synaptic proteins (NRGN, SNAP25, ENO2) were not comparable across matrices without strict pre-analytical standardisation.

## Introduction

Blood-based biomarkers now inform the diagnosis and prognostic stratification of neurodegenerative disease. Plasma phosphorylated tau at threonine 217 (pTau-217), glial fibrillary acidic protein (GFAP) and neurofilament light chain (NfL) reach diagnostic accuracy comparable to cerebrospinal fluid (CSF) for Alzheimer’s disease and several neurodegenerative conditions [17, 18, 19, 20]. Beyond single-analyte assays, high-multiplex platforms such as the Olink Proximity Extension Assay, SomaScan aptamer arrays and, more recently, the NUcleic-acid-Linked Immuno-Sandwich Assay (NULISA) quantify hundreds of proteins from microlitre blood volumes [1, 8]. The NULISAseq CNS Disease Panel covers 130 proteins spanning neurodegeneration, neuroinflammation, synaptic function and amyloid processing, and has been used in cohorts ranging from Alzheimer’s disease, frontotemporal disorders and traumatic brain injury to ethnically diverse populations and remote fingerstick sampling [3, 4, 5, 6, 7, 9, 10, 11, 25].

The choice of blood matrix is one of the least examined sources of pre-analytical variability. Serum and plasma are produced differently: serum forms through complete coagulation, which releases platelet alpha-granule contents (the platelet-derived analytes classified below as Tier 3; enumerated in the Discussion), while plasma is collected with anticoagulants (EDTA or citrate) that block clotting and preserve circulating cells [22]. Even carefully processed serum and plasma are thus distinct biochemical compartments. This matters because much of the existing neurological biobank material was collected in serum-separator tubes [22], whereas most current plasma studies use K2- or K3-EDTA plasma. Whenever the matrix differs, harmonisation of biomarker concentrations and clinical cut-offs across cohorts is at risk [13, 14].

For NfL, several studies report high serum-plasma concordance, with both matrices giving clinically equivalent information after calibration [13, 14, 15]. Plasma is generally treated as the reference matrix for phosphorylated tau, yet recent work suggests that serum NULISA captures much of the same dynamic range with a systematic offset [2, 22]. Only one preliminary evaluation of serum as an alternative NULISA matrix has been published [2], and it covered a limited subset of proteins in a smaller cohort. The full NULISAseq CNS Disease Panel has not been compared systematically across serum and plasma.

We therefore performed the first complete head-to-head evaluation of all 130 proteins of the NULISAseq CNS Disease Panel in 62 paired serum and plasma samples from a clinical neurological cohort spanning Alzheimer’s disease, non-Alzheimer’s dementias, motor neuron disease, multiple sclerosis and healthy controls. The aims were to (i) quantify rank and absolute concordance for each of the 130 analytes, (ii) identify proteins for which serum and plasma are interchangeable in clinical use, and (iii) define proteins for which systematic matrix biases preclude direct comparison across cohorts.

## Methods

### Study population and design

This was a single-centre, evaluation in which each participant donated serum and plasma simultaneously at the same venipuncture, ensuring a within-subject paired design that eliminates inter-individual variability from the concordance estimate. The study was conducted at Spedali Civili di Brescia, Italy. Participants were drawn from the same cohorts previously reported, from which the present paired serum and plasma samples originate (Pilotto et al. [33]; Martinuzzo et al. [34]). No randomisation was performed: this was a within-subject method-comparison study without group allocation or intervention. Participant-level blinding was not applicable; NULISA measurements were obtained by laboratory personnel blinded to clinical diagnosis through anonymised sample identifiers. Eligible participants were consecutive individuals attending the centre with a clinically established diagnosis (Alzheimer’s disease, non-Alzheimer’s dementia, motor neuron disease or multiple sclerosis) or healthy control status, for whom serum and plasma had been collected simultaneously at the same venipuncture. Participants were excluded if a matched serum-plasma pair was unavailable or if either aliquot failed pre-analytical quality control. Diagnoses followed standard criteria: Alzheimer’s disease, NIA-AA criteria [27]; multiple sclerosis, 2024 revisions of the McDonald criteria [28]; motor neuron disease, revised El Escorial [29] and Gold Coast [30] criteria; within the non-Alzheimer’s dementia group, behavioural-variant frontotemporal dementia [31] and dementia with Lewy bodies [32].

The study was approved by the local ethics committee (DMA study, NP 1471; and Neuromultibio study, NP 5285, approved by the local Brescia ASST ethics committee on 10.05.2022) and was performed in conformity with the Helsinki Declaration; informed consent was obtained from each participant or their legal representative. The final analytical cohort comprised 62 participants (mean age 56.4 +/- 19.7 years, range 20-84 years; 39 (62.9%) female) including 30 with neurodegenerative conditions, 19 with multiple sclerosis and 13 healthy controls (see Results and Table 1 for details). Age and sex differed across diagnostic groups, with the multiple sclerosis group younger (mean 35.1 years) and the Alzheimer’s disease and non-Alzheimer’s dementia groups older (70.5 and 70.1 years). Because each participant contributed serum and plasma from the same venipuncture, this paired within-subject design removes age, sex and any subject-level comorbidity as confounders of the concordance estimates; age and sex were recorded and must be weighed in any interpretation tied to clinical diagnosis.

**Table 1.** Cohort characteristics.

| Characteristic | Total (n=62) | AD (n=18) | HC (n=13) | MND (n=4) | NAD (n=8) | MS (n=19) |
| --- | --- | --- | --- | --- | --- | --- |
| Age, mean ± SD (years) | 56.4 ± 19.7 | 70.5 ± 7.6 | 57.4 ± 17.0 | 63.2 ± 7.1 | 70.1 ± 9.4 | 35.1 ± 15.8 |
| Female, n (%) | 39 (62.9%) | 7 (38.9%) | 12 (92.3%) | 3 (75.0%) | 4 (50.0%) | 13 (68.4%) |
| Male, n (%) | 23 (37.1%) | 11 (61.1%) | 1 (7.7%) | 1 (25.0%) | 4 (50.0%) | 6 (31.6%) |

*Legend. AD = Alzheimer's disease; HC = healthy controls; MND = motor neuron disease; MS = multiple sclerosis; NAD = non-Alzheimer's dementia. Values are mean ± standard deviation or n (%).*

**Table 2.**
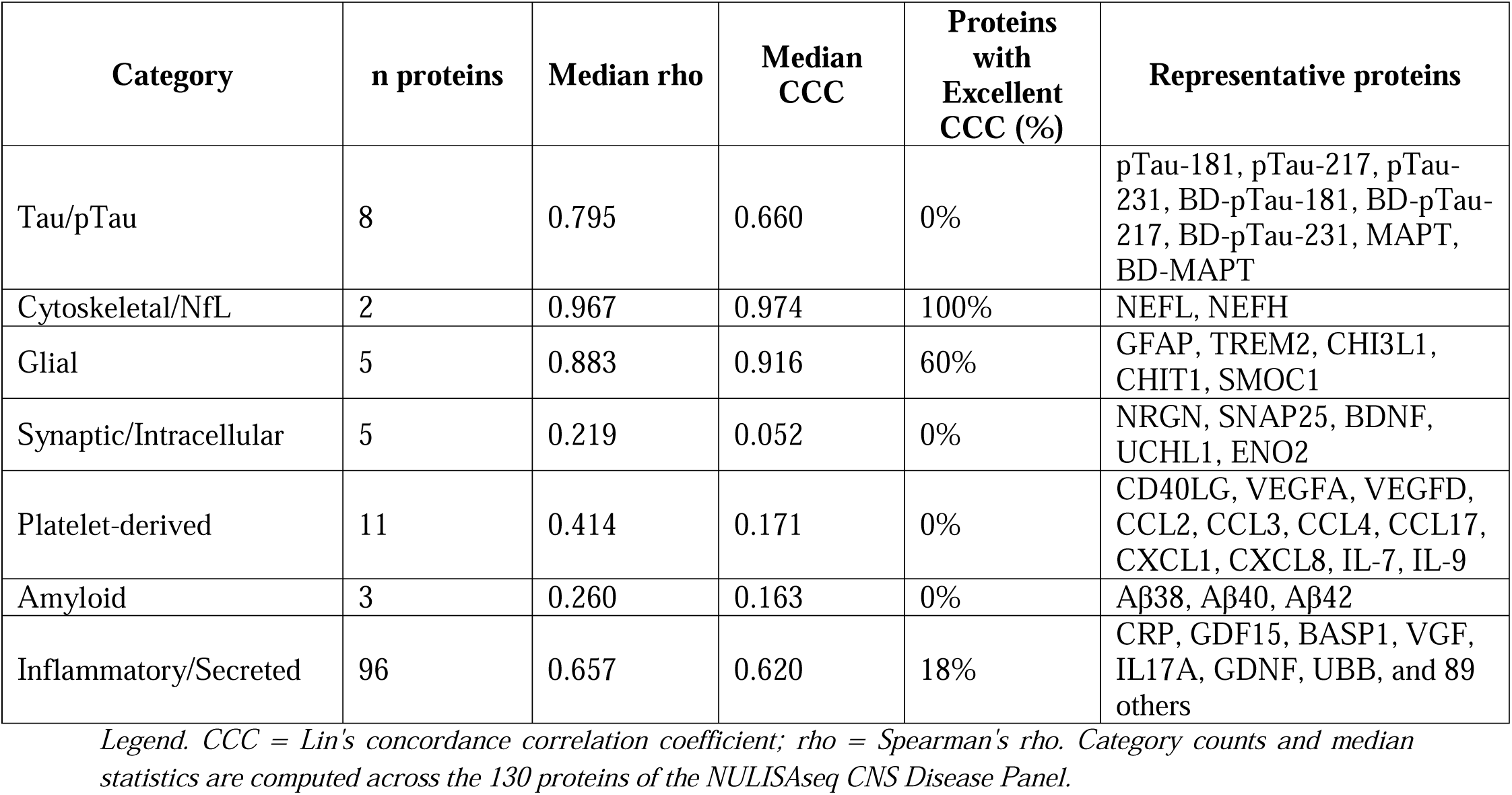
Concordance summary by protein category.

**Table 3.** Top 10 most concordant and 10 least concordant proteins.

| Protein | Spearman rho<br>[95% CI] | CCC | ICC | dNPQ | Category |
| --- | --- | --- | --- | --- | --- |
| Top 10 - most concordant |  |  |  |  |  |
| GDNF | 0.988 | 0.990 | - | +0.21 | Other |
| NEFH | 0.988 | 0.988 | 0.988 | -0.164 | Cytoskeletal |
| UBB | 0.941 | 0.990 | - | +0.15 | Housekeeping |
| BASP1 | 0.972 | 0.986 | - | +0.12 | Other |
| IL17A | 0.961 | 0.977 | - | +0.10 | Inflammatory |
| VGF | 0.953 | 0.980 | - | +0.09 | Other |
| PDLIM5 | 0.949 | 0.982 | - | +0.05 | Other |
| GFAP | 0.949 | 0.939 | 0.940 | +0.010 | Glial |
| NEFL | 0.947 | 0.960 | 0.960 | -0.051 | Cytoskeletal |
| GDF15 | 0.941 | 0.918 | - | +0.53 | Secreted |
| Bottom 10 - least concordant |  |  |  |  |  |
| Abeta38 | 0.010 | -0.030 | - | - | Amyloid |
| SOD1 | 0.073 | 0.087 | - | -0.79 | Other |
| SFRP1 | 0.070 | 0.039 | - | +1.47 | Other |
| CD40LG | 0.102 | 0.003 | 0.003 | -4.74 | Platelet-derived |
| IL7 | NS | 0.031 | - | -2.41 | Inflammatory |
| BDNF | 0.223 | 0.027 | - | -2.69 | Platelet-derived |
| ENO2 | NS | 0.019 | - | +5.72 | Intracellular |
| SNAP25 | 0.219 | 0.078 | - | NS | Synaptic |
| NRGN | 0.147 | 0.053 | - | +4.39 | Synaptic |
| VEGFA | 0.168 | 0.048 | - | -0.92 | Platelet-derived |

*Legend. CCC = Lin's concordance correlation coefficient; ICC = intraclass correlation coefficient (2,1); dNPQ = plasma - serum mean paired difference in NULISA Protein Quantification units; NS = not significant after FDR correction; '-' = not reported.*

### Blood collection and sample processing

At each study visit, one serum-separator tube (SST; 4 mL) and one K2-ethylenediaminetetraacetic acid (K2-EDTA; 4 mL) tube were collected simultaneously at a single venipuncture, with the SST drawn before the K2-EDTA tube in accordance with the standard order of draw. After collection, SST tubes were left to clot at room temperature for 30 minutes before centrifugation; K2-EDTA tubes were gently inverted five to ten times immediately after collection to ensure uniform anticoagulant mixing. Both tube types were centrifuged at 2500×g for 10 minutes at room temperature. Serum and plasma aliquots (0.5 mL each) were pipetted into polypropylene cryotubes and stored at −80°C until analysis. All samples underwent a single freeze-thaw cycle prior to the NULISA run. On the day of analysis, aliquots were thawed at room temperature (21–23°C) and centrifuged at 10000×g for 5 minutes immediately before loading.

Samples were processed following current recommendations for pre-analytical standardisation of blood biomarkers [22].

### NULISA proteomic profiling

NULISA assays were performed at the Neurobiorepository and Laboratory of Advanced Biological Markers, University of Brescia and ASST Spedali Civili Hospital (Brescia, Italy). Serum and plasma samples were processed across four plates (plate runs: 25/09/2025, 01/10/2025, 09/10/2025, 29/10/2025) across two reagent lots (lot b2: n=42; lot b3: n=20). For the NULISAseq CNS Disease Panel, 130 neurodegenerative disease-related proteins were quantified through immunocomplex formation associated with DNA reporter molecule ligation. DNA reporter molecules were pooled, amplified by PCR, purified and sequenced on the Illumina NextSeq 2000 platform. Sequencing data were processed using the NULISAseq algorithm (Alamar Biosciences, Newark, CA, USA). Intraplate normalisation was performed by dividing the target counts for each sample well by that well’s internal control counts; interplate normalisation was then applied using interplate control (IPC) normalisation, whereby counts were divided by target-specific medians of the three IPC wells on each plate. Data were rescaled and log2-transformed to obtain NULISA Protein Quantification (NPQ) units for downstream statistical analysis. All sample measurements with NPQ values below the limit of detection (LOD), as provided by Alamar Biosciences, were excluded from per-protein analyses. Research resources: NULISAseq CNS Disease Panel and Alamar ARGO instrument (Alamar Biosciences, Newark, CA, USA); Illumina NextSeq 2000 (Illumina, San Diego, CA, USA). NULISA CNS Panel/kit’s catalogue number is as follows: 800104.

The panel comprised 130 proteins spanning neurodegeneration markers (pTau-181, pTau-217, pTau-231, GFAP, NEFL, NEFH), microglial and glial markers (TREM2, CHI3L1, CHIT1), inflammatory cytokines and chemokines, synaptic proteins (NRGN, SNAP25, BDNF) and amyloid peptides (Abeta38, Abeta40, Abeta42). The panel additionally includes brain-derived phosphorylated tau variants designated as BD-pTau-181, BD-pTau-217, BD-pTau-231 and BD-MAPT, which use antibody pairs targeting brain-enriched phospho-epitopes distinct from the standard pTau assay configurations; full antibody specifications are available in the Alamar NULISAseq CNS Disease Panel technical documentation. Protein abundance was expressed in NULISA Protein Quantification (NPQ) units, i.e., log2-normalised counts derived from the NULISAseq pipeline [1, 3].

### Inter-batch bridging

Samples were processed across two reagent lots (lot b2: n=42; lot b3: n=20). The primary concordance analysis used non-bridged (raw). NPQ values from within-lot matched serum-plasma pairs, ensuring that no confounding inter-lot correction could artificially inflate or deflate the matrix-comparison signal. Post-bridging values were retained for a pre-specified sensitivity analysis. Inter-lot bridging was implemented using an Olink-derived workflow adapted for NULISA NPQ data. Lot b2 (n = 42) was designated as the reference dataset; bridge samples were selected from lot b2 on the basis of satisfactory quality-control metrics, high target detectability, a low proportion of values below the limit of detection, and representativeness of the full analytical dynamic range as confirmed by principal component analysis. For each analyte, a per-protein adjustment factor was computed as the median pairwise NPQ difference between lot b2 and lot b3 values across the bridge sample set, and this factor was applied additively to all lot b3 NPQ values to project them onto the b2 reference scale. Bridge quality was evaluated per analyte using five metrics: median NPQ difference before and after bridging, mean absolute deviation (MAD), Spearman’s rank correlation, and the slope of the linear regression X = a + b × X. Analytes failing predefined quality thresholds were flagged and excluded from the bridged dataset retained for sensitivity analyses. All bridging procedures were performed in R (version 4.4.2).

### Statistical analysis

All statistical analyses were performed in R version 4.5.1 (2025-06-13) (R Foundation for Statistical Computing, Vienna, Austria). Key package versions: epiR 2.0.93, psych 2.6.5, broom 1.0.10, ggplot2 4.0.1, dplyr 1.1.4, tidyverse 2.0.0. For each of the 130 proteins, rank concordance between serum and plasma was quantified using Spearman’s rho with 95% confidence intervals derived by Fisher z-transformation. Multiple testing was controlled using the Benjamini-Hochberg false discovery rate (FDR), with the adjusted p-value reported as padj. Lin’s concordance correlation coefficient (CCC) was calculated using the epiR package (function epi.ccc()), with 95% CI obtained by bootstrap (1000 resamples) [16]. CCC values were categorised as Excellent (CCC >= 0.90), Good (0.80 <= CCC < 0.90), Moderate (0.65 <= CCC < 0.80) and Poor (CCC < 0.65); these thresholds were adopted operationally for this study, analogous to those used in prior method-comparison analyses. Intraclass correlation coefficient (ICC(2,1); two-way random effects, absolute agreement) was computed using the psych package. Matrix-specific differences were quantified as the mean paired difference in NPQ (dNPQ = plasma - serum) and tested with two-tailed paired t-tests; because NPQ units are log2-transformed, paired differences correspond to log-ratios. Normality of the per-protein paired differences was assessed with the Shapiro-Wilk test; the null of normality was rejected (p < 0.05) for 51 of 130 proteins (43 after Benjamini-Hochberg correction). All paired comparisons were therefore repeated with the distribution-free Wilcoxon signed-rank test as a sensitivity analysis: the parametric and non-parametric tests agreed on FDR significance for 128 of 130 proteins (98.5%), with no protein reaching significance under the t-test alone, indicating that the dNPQ findings are robust to departures from normality; the rank-based and agreement metrics (Spearman’s rho, CCC, ICC) do not assume normality. Per-protein Shapiro-Wilk and Wilcoxon results are reported in Supplementary Table S1. No outlier exclusion or winsorisation was performed: all measurements above the LOD were retained and no formal outlier test was applied. Per-protein valid pair counts (both matrices above LOD), t statistics and degrees of freedom, with exact p-values and padj, are reported in Supplementary Table S1. Significance thresholds for the volcano analysis were |dNPQ| >= 0.5 and padj < 0.05. All concordance analyses were repeated on the bridged dataset as a sensitivity analysis. No formal a priori sample-size calculation was performed: this was a within-subject method-comparison study in which all participants with simultaneously collected serum and plasma were included. The resulting sample of 62 paired specimens affords adequate precision for the agreement estimates — under Fisher z-transformation, the two-sided 95% confidence interval for a concordance/correlation coefficient of 0.80 has a half-width of approximately 0.09, narrowing to ≤0.05 for coefficients ≥0.90 — and is in line with established recommendations for reliability and method-comparison studies.

## Results

### Cohort characteristics

Sixty-two individuals with paired serum and plasma samples were included (Table 1). The cohort had a mean age of 56.4 +/- 19.7 years (range 20-84 years), with 39 (62.9%) women and 23 (37.1%) men, including 30 neurodegenerative conditions, 19 demyelinating diseases and 13 controls (Table1).

### Global concordance across 130 proteins (Figure 1 and Figure 2)

Across the full panel of 130 proteins, the median Spearman rho between serum and plasma was approximately 0.72, with values ranging from 0.10 to 0.988. After Benjamini-Hochberg correction, 123 of 130 proteins (94.6%) reached FDR significance at padj < 0.05, indicating that the majority of analytes carry shared rank information between the two matrices. Eighteen proteins achieved rho >= 0.90. IL12p70 (rho=0.895) approached but did not reach the 0.90 threshold and was therefore excluded from this count (it is reported in the near-threshold group in Supplementary Table S1). By design, the within-subject paired estimate of concordance is not confounded by age, sex, comorbidity or diagnosis, since both matrices derive from the same individual and venipuncture. Conversely, four proteins were non-concordant by rank and failed to reach FDR significance: CD40LG (rho=0.102, padj=0.44), SOD1 (rho=0.073, padj=0.58), SFRP1 (rho=0.070, padj=0.59) and Abeta38 (rho=0.010, padj=0.94). Within the core neurodegeneration markers, pTau-181 (rho=0.869, 95% CI 0.79-0.92, padj=2.7e-19), pTau-217 (rho=0.846, 95% CI 0.76-0.90, padj=2.3e-17), pTau-231 (rho=0.885, 95% CI 0.81-0.93, padj=9.7e-21), GFAP (rho=0.949, padj=1.8e-30), NEFL (rho=0.947, padj=4.0e-30), NEFH (rho=0.988, padj=1.6e-48), TREM2 (rho=0.889, padj=3.4e-21) and CHI3L1 (rho=0.877, padj=5.3e-20) all showed high rank concordance.

**Figure 1.**
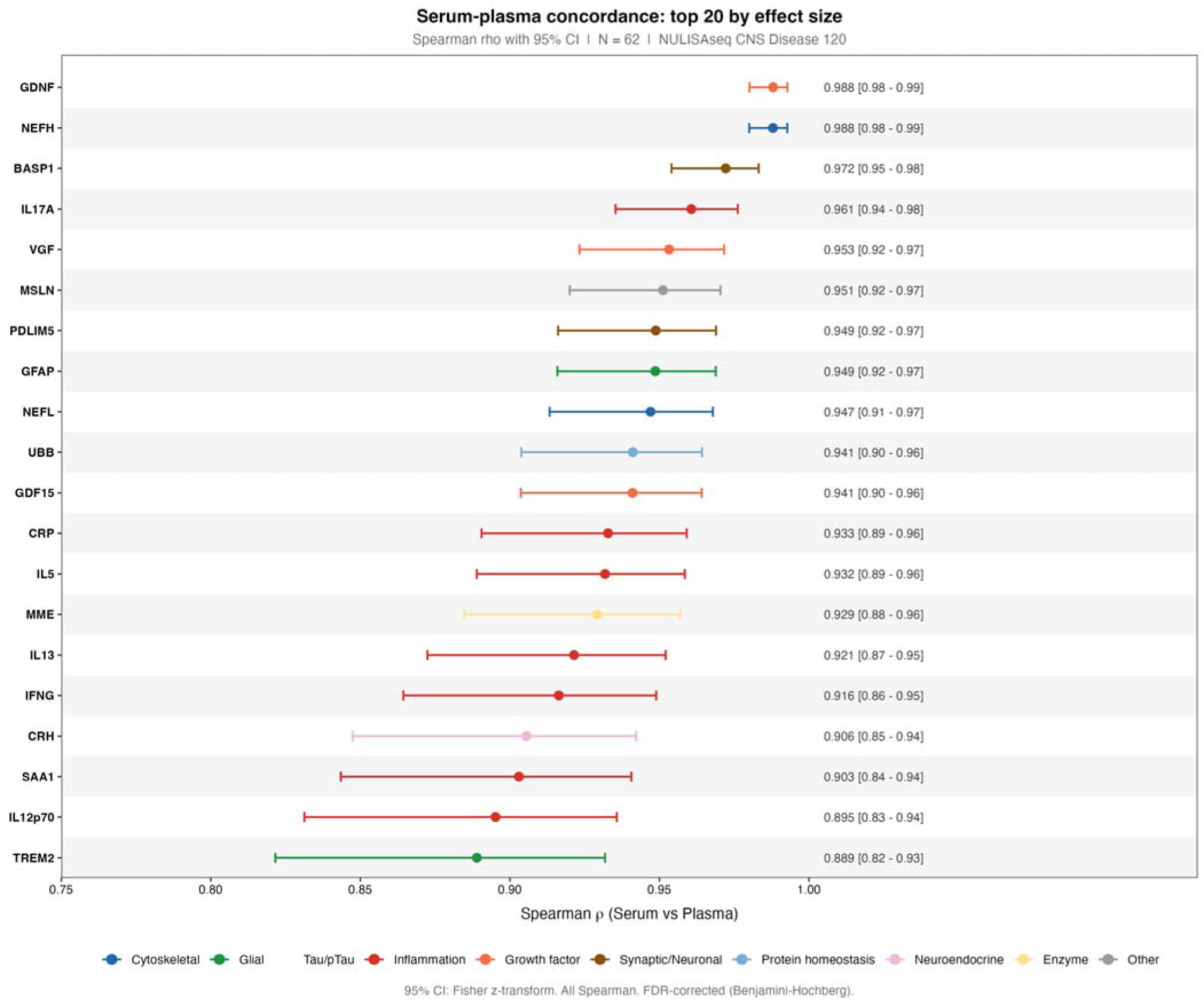
Forest plot of Spearman’s rho — top 20 most concordant proteins. Forest plot showing Spearman’s rho with 95% confidence intervals (Fisher z-transformation) for the 20 proteins with highest rank concordance across the NULISAseq CNS Disease Panel (n = 62 paired serum-plasma samples, primary non-bridged analysis). Each point represents the Spearman rho estimate; horizontal bars indicate the 95% CI. The dashed vertical line at rho = 0.90 marks the ’Excellent’ concordance threshold. FDR significance is indicated by asterisks (* padj < 0.05, ** padj < 0.01, *** padj < 0.001). Proteins are coloured by functional class.

**Figure 2.**
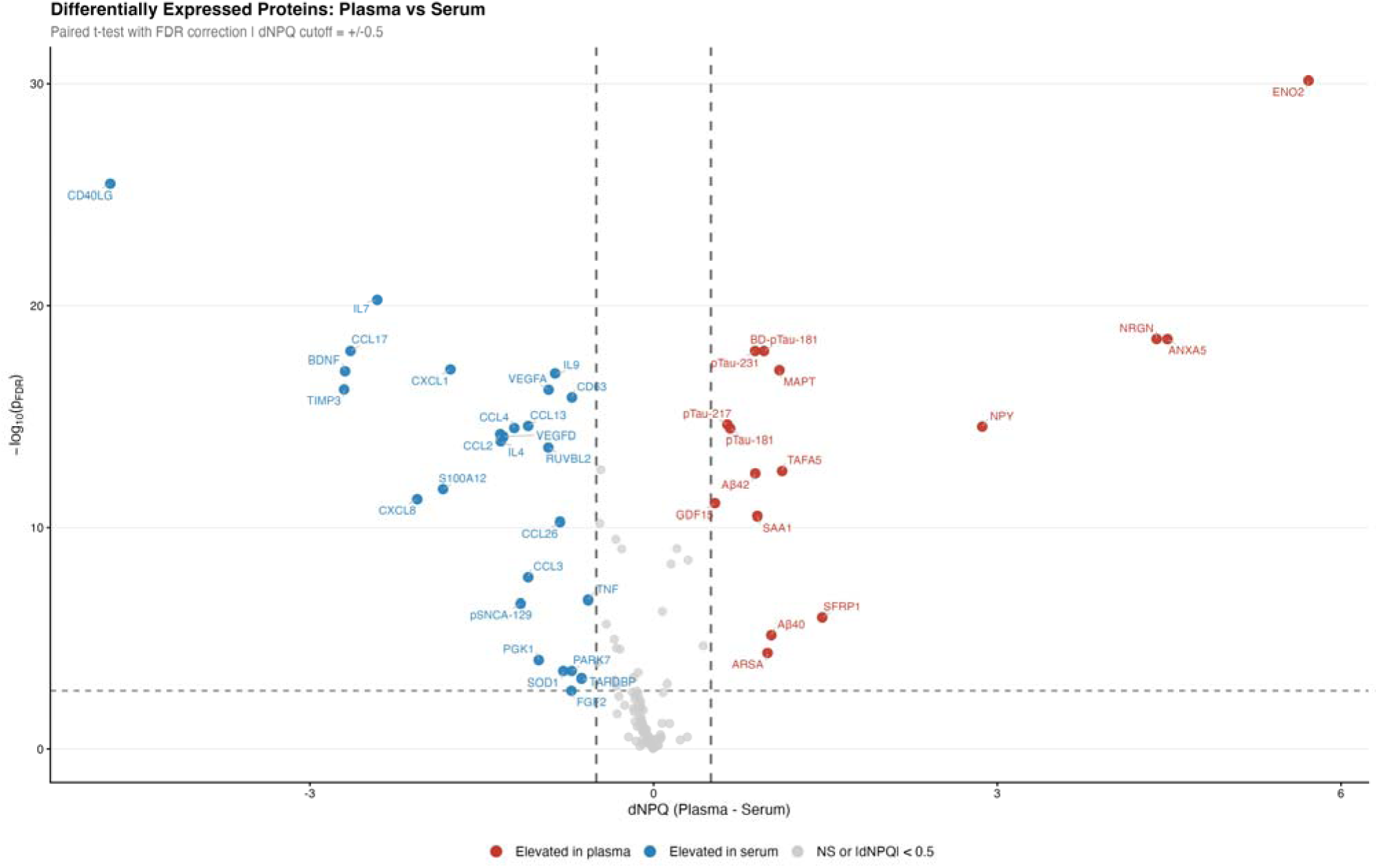
Matrix-specific bias: dNPQ volcano plot. Volcano plot of the mean paired plasma minus serum difference (dNPQ = plasma − serum, NPQ units) on the x-axis against -log10(padj) on the y-axis, for each of the 130 proteins of the NULISAseq CNS Disease Panel (primary non-bridged analysis, n = 62 pairs). Vertical dashed lines at dNPQ = -0.5 and +0.5 NPQ define the bias thresholds; the horizontal dashed line marks padj = 0.05. Plasma-enriched proteins (dNPQ > +0.5, padj < 0.05) cluster on the right and include intracellular and synaptic markers (ENO2, ANXA5, NRGN, MAPT, pTau-181, pTau-217, pTau-231, Abeta40, Abeta42). Serum-enriched proteins (dNPQ < -0.5, padj < 0.05) cluster on the left and are dominated by platelet-derived and leukocyte-derived analytes (CD40LG, BDNF, CCL17, IL-7, CXCL8, CCL2/3/4, VEGFA/D).

### Absolute agreement: Lin’s CCC and ICC

The CCC and ICC assessment across the 130 proteins indicated 22 proteins reaching the ’Excellent’ category (CCC >= 0.90) and further 13 proteins reached ’Good’ agreement (0.80 <= CCC < 0.90). Approximately 50 proteins fell into the ’Moderate’ band (0.65-0.80) and 38 into ’Poor’ (<0.65) (the remaining proteins were not classifiable owing to insufficient valid pairs).

The contrast between rho and CCC was most informative for analytes with high rank concordance but a systematic NPQ offset. The phosphorylated tau species illustrate this pattern: pTau-181 had rho=0.869 but CCC=0.717 (Moderate), pTau-217 had rho=0.846 but CCC=0.699, and pTau-231 had rho=0.885 but CCC=0.679. The corresponding ICC (2,1) values (pTau-181 ICC=0.720, pTau-217 ICC=0.703, pTau-231 ICC=0.682) closely mirrored the CCC, confirming that the loss of agreement was driven by a location shift rather than by random scatter. In contrast, GFAP (CCC=0.939, ICC=0.940) and NEFL (CCC=0.960, ICC=0.960) showed near-perfect agreement on both indices. At the opposite extreme, CD40LG showed essentially nihil agreement on all three statistics (rho=0.102, CCC=0.003, ICC=0.003). Full per-protein values are reported in Supplementary Table S1.

### Systematic matrix biases: dNPQ volcano analysis (Figure 2)

To characterise the direction and magnitude of matrix-specific bias, we computed the mean paired difference dNPQ = plasma - serum for each protein and tested it against zero using paired t-tests with FDR correction. With cut-offs of |dNPQ| >= 0.5 NPQ and padj < 0.05, two well-defined clusters emerged.

Plasma-enriched proteins (dNPQ > +0.5 NPQ) were dominated by intracellular and synaptic molecules: ENO2 (+5.72, padj=7.2e-31), ANXA5 (+4.49, padj=3.3e-19), NRGN (+4.39, padj=3.2e-19), NPY (+2.87, padj=3.0e-15), SFRP1 (+1.47, padj=1.1e-6), TAFA5 (+1.12, padj=2.9e-13), MAPT (+1.10, padj=8.0e-18), Abeta40 (+1.03, padj=7.3e-6), ARSA (+0.99, padj=4.7e-5), BD-pTau-181 (+0.96, padj=1.2e-18), SAA1 (+0.90, padj=3.1e-11), Abeta42 (+0.89, padj=3.7e-13), pTau-231 (+0.89, padj=1.2e-18), pTau-181 (+0.67, padj=3.6e-15), pTau-217 (+0.64, padj=2.4e-15) and GDF15 (+0.53, padj=8.3e-12).

Serum-enriched proteins (dNPQ < -0.5 NPQ) were dominated by platelet- and leukocyte-derived mediators, with a minority of high-abundance intracellular proteins (e.g., PGK1, SOD1, PARK7, RUVBL2, TARDBP): CD40LG (−4.74, padj=3.3e-26), TIMP3 (−2.70, padj=6.0e-17), BDNF (−2.69, padj=9.0e-18), CCL17 (−2.65, padj=1.2e-18), IL7 (−2.41, padj=5.4e-21), CXCL8 (−2.06, padj=5.5e-12), S100A12 (−1.84, padj=1.9e-12), CXCL1 (−1.77, padj=7.4e-18), CCL2 (−1.34, padj=6.5e-15), IL4 (−1.33, padj=1.3e-14), VEGFD (−1.31, padj=8.3e-15), CCL4 (−1.22, padj=3.4e-15), CCL13 (−1.09, padj=2.7e-15), CCL3 (−1.09, padj=1.8e-8), PGK1 (−1.00, padj=9.8e-5), VEGFA (−0.92, padj=6.2e-17), RUVBL2 (−0.92, padj=2.5e-14), IL9 (−0.86, padj=1.1e-17), CCL26 (−0.82, padj=5.7e-11), SOD1 (−0.79, padj=3.1e-4), CD63 (−0.71, padj=1.4e-16), PARK7 (−0.71, padj=3.1e-4), FGF2 (−0.72, padj=2.3e-3), TARDBP (−0.63, padj=6.5e-4) and TNF (−0.57, padj=1.9e-7).

A third, smaller cluster of analytes was matrix-equivalent (|dNPQ| < 0.5 and not significant after FDR correction): GFAP (dNPQ=+0.010), NEFL (−0.051), NEFH (−0.164) and CRP (−0.054). Together with their high CCC values, these proteins represent the operationally interchangeable subset of the panel.

### Class-specific concordance of key neurodegeneration biomarkers (Figure 3)

Figure 3 displays the rank concordance of 20 pre-specified clinical proteins grouped by functional class. Cytoskeletal proteins were the most matrix-stable: NEFH (rho=0.988) and NEFL (rho=0.947) had an excellent rank agreement and matrix-equivalent dNPQ (|dNPQ| < 0.2 NPQ, not significant). The glial cluster also performed extremely well: GFAP (rho=0.949, CCC=0.939, dNPQ=+0.010), TREM2 (rho=0.889), CHI3L1 (rho=0.877) and SMOC1 (rho=0.760).

**Figure 3.**
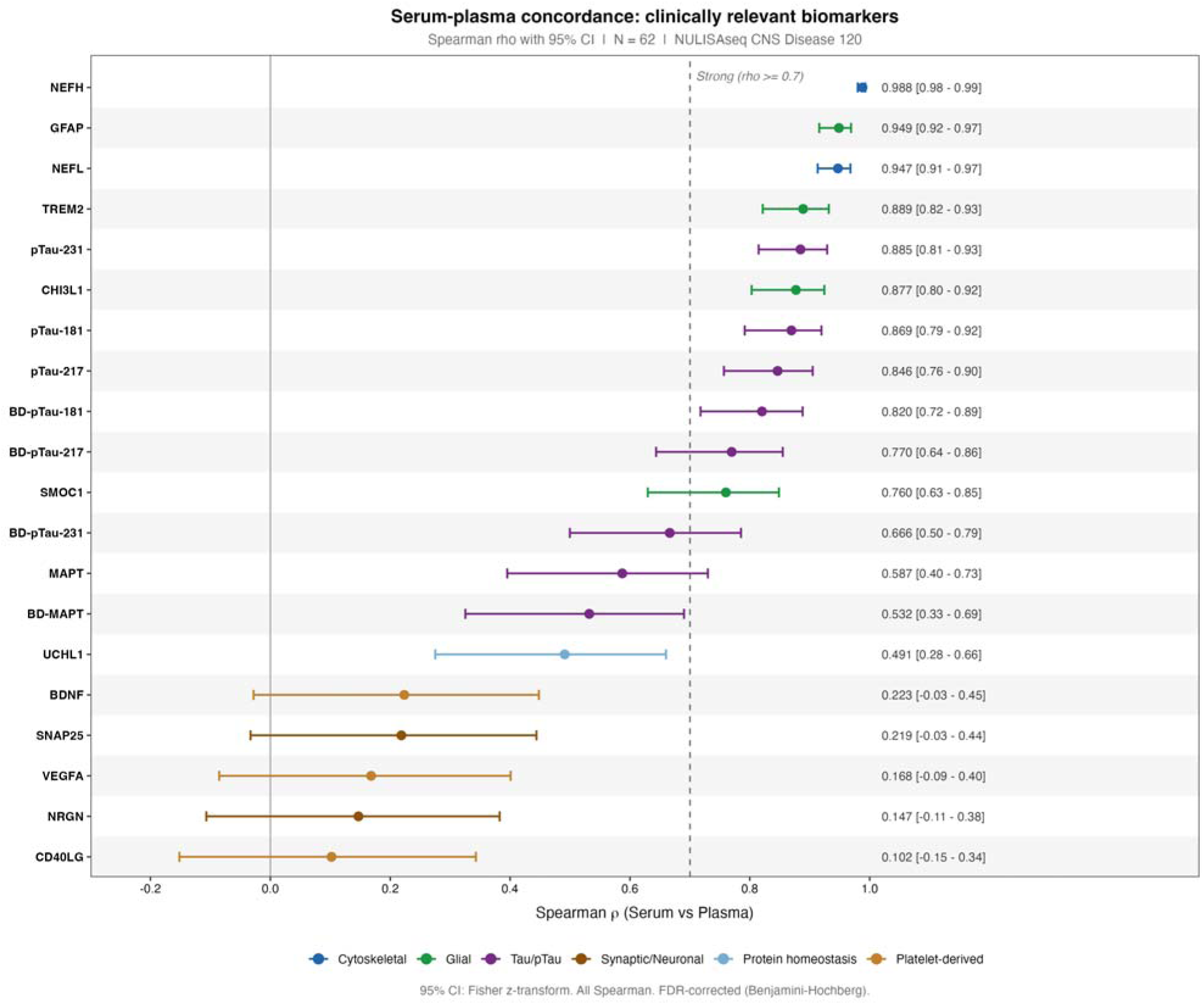
Forest plot of Spearman’s rho — 20 pre-specified clinical proteins. Forest plot showing Spearman’s rho with 95% confidence intervals for 20 pre-specified neurology-relevant proteins, grouped by functional class (cytoskeletal, glial, tau/pTau, platelet-derived, synaptic/neuronal). Proteins are ordered within each class from highest to lowest rho (primary non-bridged analysis). Reference lines are placed at rho = 0.90 (Excellent), rho = 0.80 (Good) and rho = 0. The figure illustrates the three-tier concordance pattern: cytoskeletal and glial markers cluster near rho = 1, tau and phosphorylated tau species occupy the upper-middle range, and platelet-derived and synaptic proteins cluster near rho = 0.

Tau and phosphorylated tau species formed a distinct intermediate class. Their rank concordance was excellent (pTau-181 rho=0.869, pTau-217 rho=0.846, pTau-231 rho=0.885, BD-pTau-181 rho=0.820, BD-pTau-217 rho=0.770, BD-pTau-231 rho=0.666, MAPT rho=0.587, BD-MAPT rho=0.532), but in each case there was a systematic plasma-greater-than-serum offset of approximately +0.64 to +0.89 NPQ units. Consequently, CCC values for phosphorylated tau fell into the Moderate band (0.679-0.770), with BD-pTau-181 (CCC=0.641) and BD-pTau-231 (CCC=0.593) reaching only the Poor category.

At the opposite end, platelet-derived and intracellular synaptic proteins were essentially non-concordant. CD40LG (rho=0.102, CCC=0.003, dNPQ=-4.74), BDNF (rho=0.223, CCC=0.027, dNPQ=-2.69), VEGFA (rho=0.168, CCC=0.048, dNPQ=-0.92), NRGN (rho=0.147, CCC=0.053, dNPQ=+4.39), SNAP25 (rho=0.219, CCC=0.078) and UCHL1 (rho=0.491) showed weak or absent rank correlation combined with very large matrix offsets, confirming that these proteins carry fundamentally different signals in serum and plasma.

### Sensitivity analysis: post-bridging data

The concordance pattern was unchanged by inter-batch correction. After applying the bridging procedure between lots b2 and b3, differences in CCC values were minimal across the panel (median delta CCC approximately -0.004), and two proteins changed their concordance category between non-bridged and bridged analyses: BD-pTau-217 showed a marginal decrease from CCC=0.760 (Good) in the non-bridged analysis to CCC=0.707 (Moderate) in the bridged analysis; IL12p70 showed a marginal decrease from CCC=0.917 (Excellent) to CCC=0.893 (Good), remaining in the high-concordance range. Both changes were small (|ΔCCC| ≤ 0.053) and did not alter the overall interpretation. The full per-protein comparison is reported in Supplementary Table S2 and Supplementary Figure S4.

## Discussion

In this paired serum-plasma comparison of the NULISAseq CNS Disease Panel, the two matrices correlated well overall. By design, the within-subject paired estimate of concordance is not confounded by age, sex, comorbidity or diagnosis, because both matrices come from the same individual and venipuncture. Concordance varied widely by protein subtype, splitting the panel into three tiers. Tier 1 holds fully interchangeable proteins, mainly cytoskeletal, glial and selected circulating cytokines (see Supplementary Table S1). Tier 2 holds rank-concordant but offset-shifted proteins, typified by phosphorylated tau species and amyloid peptides, where rho stays high (>= 0.80) but a systematic plasma-greater-than-serum location shift pushes CCC into the Moderate band. Tier 3 holds matrix-dependent proteins, dominated by platelet-derived analytes (CD40LG, BDNF, VEGFA, CCL2/3/4/17, IL-7, IL-9, CXCL1/8) and intracellular synaptic proteins (NRGN, SNAP25, ENO2), where rho is close to zero and CCC near null. The same tier structure holds across rho, CCC, ICC and dNPQ, and tells investigators whether serum and plasma results can be pooled within or across studies.

During coagulation, platelets activate and empty their alpha-granules into the forming serum, releasing CD40 ligand (sCD40L), BDNF, VEGFA, VEGFD, PF4, CXCL chemokines, IL-7 and several CCL chemokines [22]. These analytes therefore run systematically higher in serum than in EDTA plasma. Clotting can also sequester or degrade other proteins. ENO2 (gamma-enolase, also called neuron-specific enolase, NSE), expressed mainly in neurons and neuroendocrine cells and only weakly in erythrocytes and platelets, was markedly higher in EDTA plasma, in keeping with protection from coagulation-driven adsorption or degradation during clot formation. NRGN (neurogranin), a postsynaptic calmodulin-binding protein of neuronal origin, was likewise raised in plasma, where intracellular neuronal contents are better preserved under anticoagulation. The direction of the matrix bias thus tracks a protein’s relative platelet and blood-cell abundance: platelet and cell lysis during clotting pushes high-abundance intracellular proteins into serum (as seen for SOD1 and PGK1), whereas for the low-platelet, mainly neuronal proteins ENO2 and NRGN, adsorption and degradation within the clot dominate and leave them enriched in plasma. ANXA5 (Annexin A5, dNPQ = +4.49), a phosphatidylserine-binding protein of platelets and vascular endothelium, was also unexpectedly higher in plasma; the likeliest reason is partial trapping within the fibrin clot during serum formation, which lowers its serum concentration relative to anticoagulated plasma. Phosphorylated tau and amyloid peptides may bind plasma proteins or undergo differential proteolysis during clotting, in line with the modest but reproducible plasma-greater-than-serum offset we saw for pTau-181, pTau-217, pTau-231, Abeta40 and Abeta42 [22]. Two proteins normally read as markers of erythrocyte contamination behaved unexpectedly. Farinas et al. [2] applied the erythrocyte signature of Geyer et al. [26] (HBA1, PRDX6, SOD1, PGK1) and found all four higher in plasma; our data split that panel. SOD1 (dNPQ = -0.79, padj = 3.1e-4) and PGK1 (dNPQ = -1.00, padj = 9.8e-5) were higher in serum, while HBA1 (dNPQ = +0.23) and PRDX6 (dNPQ = -0.22) did not differ between matrices. SOD1 and PGK1 are abundant in platelets as well as red cells, so platelet activation during clotting drives them into serum alongside the other Tier 3 platelet analytes (CD40LG, BDNF, VEGFA, the CCL chemokines). The discordance with Farinas et al. probably reflects pre-analytical conditions: their tubes were centrifuged at 4 degrees C, against our 2500xg at room temperature, which favours platelet activation. For platelet-sensitive analytes the very direction of the bias depends on the protocol, a further reason they cannot be compared across matrices or cohorts without strict pre-analytical standardisation.

These findings line up with the existing serum-plasma literature and widen it. For NfL, our CCC values for NEFL (0.960) and NEFH (0.988) match earlier serum-plasma comparisons on single-molecule array (Simoa) and Lumipulse platforms [13, 14, 15], confirming that neurofilaments behave as matrix-agnostic biomarkers at current analytical resolution. The plasma-greater-than-serum offset we found for phosphorylated tau species agrees with the one previous preliminary NULISA serum evaluation [2] and with Simoa- and Lumipulse-based reports for pTau-217 and pTau-181 [20, 21, 22]. The near-complete dissociation between serum and plasma for CD40LG, BDNF and the CCL/CXCL chemokines reproduces a familiar pre-analytical signature of platelet activation [22]. What sets this study apart is its breadth: it covers the entire 130-protein NULISAseq CNS Disease Panel in one paired cohort, where earlier work was limited to small panels or single analytes. Cross-platform comparison of NULISA and Olink in a critically ill COVID-19 cohort shows broadly similar signal for shared analytes [8], suggesting that matrix-specific patterns are partly platform-independent, though this needs replication in neurological cohorts.

These results bear directly on biomarker use and on the reading of legacy biobanks. Centres holding mostly serum can use NULISA serum without conversion factors for cytoskeletal markers (NEFL, NEFH), glial markers (GFAP, TREM2, CHI3L1, CHIT1) and most circulating inflammatory mediators with CCC >= 0.90. For tau, phosphorylated tau and amyloid peptides, rank-based algorithms such as amyloid probability scores or Youden-optimal classifiers should transfer between matrices, since rho stays high, but absolute concentration thresholds must be set per matrix. For the platelet-derived Tier 3 analytes, no cross-matrix comparison is defensible, and clinical or research readings must be tied to the matrix and to standardised collection conditions [22].

The study has clear strengths. Each participant gave serum and plasma at one venipuncture, the cohort spans neurodegenerative and demyelinating disease, and four metrics (Spearman rho, CCC, ICC and dNPQ) point to the same tier structure. The 62 paired samples give adequate power for the primary concordance estimates. Limits remain. The cohort is single-centre and needs external validation in independent multi-centre datasets. Generalisability across ethnic groups is untested, given known ethnic variation in blood biomarker levels [24]. The cross-sectional design says nothing about the longitudinal stability of matrix-specific offsets. Age and sex were unevenly spread across diagnostic groups, with the multiple sclerosis group younger and the dementia groups older; the paired design stops this from biasing the concordance metrics but limits any diagnosis-level reading. Inter-lot bridging used a subset of samples, and although the sensitivity analysis was reassuring, larger multi-centre replication would strengthen the three-tier classification.

In sum, NULISA serum is a workable alternative to plasma for most clinically relevant neurodegeneration markers, leaving a defined set of analyte classes that still need matrix-specific calibration or strict pre-analytical control. Reporting matrix type, anticoagulant, collection-to-processing time and freeze-thaw history, in line with the Global Biomarker Standardization Consortium recommendations [22], remains essential for cross-cohort harmonisation. Sensible next steps are empirical conversion factors for phosphorylated tau and amyloid peptides across matched serum and plasma, validation of the three-tier classification in independent cohorts with CSF and PET confirmation [12, 23], and extension to remote sampling formats such as capillary blood and dried blood spots, where pre-analytical conditions differ further [25].

## Conclusion

Serum-plasma concordance across the 130 CNS-relevant proteins of the NULISAseq CNS Disease Panel is strongly protein-dependent. Cytoskeletal and glial markers, including NEFL, NEFH, GFAP and TREM2, showed excellent absolute agreement and suit inter-matrix comparison without conversion. Phosphorylated tau species kept excellent rank concordance but carried a systematic plasma-greater-than-serum offset, so their clinical thresholds must be matrix-specific. The platelet-derived Tier 3 analytes showed near-zero concordance and cannot be compared across matrices. Together these results give a practical basis for using NULISA data from existing serum biobanks and for designing future multi-centre studies in neurodegeneration and neuroinflammation.

## Data Availability

All data produced in the present study are available upon reasonable request to the authors. Individual-level proteomic data cannot be shared publicly because of patient-privacy regulations; aggregated concordance statistics and the R analysis code are available from the corresponding author upon reasonable request.

## Acknowledgements

APi has been supported by grants of Airalzh Foundation AGYR2021 Life-Bio Grant, The LIMPE-DISMOV Foundation Segala Grant 2021, the Italian Ministry of University and Research PRIN COCOON (2017MYJ5TH), PRIN 2022PNJS5Z and PRIn PNRR (P20224ZHM9), DIGI-BRAIN the H2020 IMI IDEA-FAST (ID853981), Italian Ministry of Health, Grant/Award Number: RF-2018-12366209, PNRR-Health PNRR-MAD-2022-12376110 and PNRR-MCNT2-2023-12378387, The MJFF Foundation Grant 022343, #NEXTGENERATIONEU (NGEU) funded by the Ministry of University and Research (MUR), National Recovery and Resilience Plan (NRRP), project MNESYS (PE0000006) – a multiscale integrated approach to the study of the nervous system in health and disease (DN. 1553 11.10.2022) – subproject DIGI-BRAIN.

APa has been supported by grants of the Italian Ministry of University and Research PRIN COCOON (2017MYJ5TH) and PRIN 2021 RePlast (20202THZAW), Prin 2022 EGADi (P2022TKN8C) the H2020 IMI IDEA-FAST (ID853981), #NEXTGENERATIONEU (NGEU) funded by the Ministry of University and Research (MUR), National Recovery and Resilience Plan (NRRP), project MNESYS (PE0000006) – a multiscale integrated approach to the study of the nervous system in health and disease (DN. 1553 11.10.2022) – subproject DIGI-BRAIN Grant/Award Number: RF-2018-12366209, RF-2019-12369272 and PNRR-Health PNRR-MAD-2022-12376110, the Next Generation EU - NRRP M6C2 - Investment 2.1 Enhancement and strengthening of biomedical research in the NHS-PNRR – PNRR PNRR-Health PNRR-MAD-2022-12376110..

## Data Availability

Data Availability: Individual-level proteomic data cannot be shared publicly due to applicable patient privacy regulations. Aggregated concordance statistics and R analysis code are available from the corresponding author upon reasonable request.

## Abbreviations

AD: Alzheimer’s disease
ANXA5: annexin A5
BD-pTau: brain-derived phosphorylated tau
CCC: Lin’s concordance correlation coefficient
CI: confidence interval
CNS: central nervous system
COI: conflict of interest
CSF: cerebrospinal fluid
dNPQ: mean paired plasma–serum difference in NPQ units
EDTA: ethylenediaminetetraacetic acid
FDR: false discovery rate
GBSC: Global Biomarker Standardization Consortium
GFAP: glial fibrillary acidic protein
HC: healthy controls
ICC: intraclass correlation coefficient
IPC: interplate control
LOD: limit of detection
MAD: mean absolute deviation
MND: motor neuron disease
MS: multiple sclerosis
NAD: non-Alzheimer’s dementia
NEFH: neurofilament heavy chain
NEFL / NfL: neurofilament light chain
NPQ: NULISA Protein Quantification
NULISA: NUcleic-acid-Linked Immuno-Sandwich Assay
PCA: principal component analysis
PCR: polymerase chain reaction
PET: positron emission tomography
pTau: phosphorylated tau
rho: Spearman’s rank correlation coefficient
SD: standard deviation
SST: serum-separator tube.

## References

[1] Feng W, et al. NULISA: a proteomic liquid biopsy platform with attomolar sensitivity and high multiplexing. Nat Commun. 2023. PMID: 37945559.

[2] Farinas MF, et al. Evaluation of serum as an alternative matrix to plasma for NULISA-based proteomic blood biomarker measurements. Sci Rep. 2026;16(1). PMID: 42000813. DOI: 10.1038/s41598-026-46409-w.

[3] Zeng X, et al. Novel plasma biomarkers of amyloid plaque pathology and cortical thickness: evaluation of the NULISA targeted proteomic platform in an ethnically diverse cohort. Alzheimers Dement. 2025. PMID: 39989429.

[4] Rea Reyes RE, et al. Targeted proteomic biomarker profiling using NULISA in a cohort enriched with risk for Alzheimer’s disease and related dementias. Alzheimers Dement. 2025. PMID: 40318118.

[5] Ashton NJ, et al. Biomarker discovery in Alzheimer’s and neurodegenerative diseases using nucleic acid linked immuno-sandwich assay. Alzheimers Dement. 2025. PMID: 40401628. DOI: 10.1002/alz.14621.

[6] Gong K, et al. High-sensitivity plasma proteomics reveals disease-specific signatures and predictive biomarkers of Alzheimer’s disease phenotypes in a large mixed-dementia cohort. Mol Neurodegener. 2025. PMID: 41250190. DOI: 10.1186/s13024-025-00909-x.

[7] Xu Y, et al. GPND-AI NULISA: a 15-protein AI classifier for diagnosis and co-pathology profiling across neurodegenerative diseases. Alzheimers Dement. 2026. PMID: 42050390. DOI: 10.1002/alz.71420.

[8] Taleb S, et al. Comparative analysis between Olink-PEA and Alamar-NULISA proteomic technologies applied to a critically ill COVID-19 cohort. Proteomics. 2025. PMID: 39930758.

[9] Li LM, et al. High-dimensional proteomic analysis for pathophysiological classification of traumatic brain injury. Brain. 2025. PMID: 39323289. DOI: 10.1093/brain/awae305.

[10] Li LM, et al. Systemic inflammation and its associations in acute moderate-severe traumatic brain injury: a cross-sectional study. Brain Behav Immun Health. 2026. PMID: 41623670.

[11] Graham N, et al. Midlife plasma proteomic profiles indicate altered amyloid and tau processing in former elite rugby players. J Neurol Neurosurg Psychiatry. 2026. PMID: 41047224.

[12] Lin W, et al. Abnormalities in core AD biomarkers precede inflammatory and glial markers in CSF in autosomal dominant Alzheimer’s disease. medRxiv [Preprint]. 2026. PMID: 41959774.

[13] Rübsamen N, et al. A method to combine neurofilament light measurements from blood serum and plasma in clinical and population-based studies. Front Neurol. 2022. PMID: 35775045. DOI: 10.3389/fneur.2022.894119.

[14] Tybirk L, et al. Neurofilament light chain - matrix comparison and long-term stability in frozen serum. Clin Chim Acta. 2025. PMID: 41038364.

[15] Hanin A, et al. Establishing age-stratified reference values for blood neurofilament light chain (NfL) using the Lumipulse platform. Rev Neurol. 2026. PMID: 42168010.

[16] Carrasco JL, et al. Comparison of concordance correlation coefficient estimating approaches with skewed data. J Biopharm Stat. 2007. PMID: 17613647. DOI: 10.1080/10543400701329463.

[17] Thijssen EH, et al. Plasma phosphorylated tau 217 and phosphorylated tau 181 as biomarkers in Alzheimer’s disease and frontotemporal lobar degeneration: a retrospective diagnostic performance study. Lancet Neurol. 2021. PMID: 34418401. DOI: 10.1016/S1474-4422(21)00214-3.

[18] Palmqvist S, et al. Discriminative accuracy of plasma phospho-tau217 for Alzheimer disease vs other neurodegenerative disorders. JAMA. 2020. PMID: 32722745.

[19] Ashton NJ, Brum WS, Di Molfetta G, et al. Diagnostic Accuracy of a Plasma Phosphorylated Tau 217 Immunoassay for Alzheimer Disease Pathology. JAMA Neurol. 2024;81(3):255–263. PMID: 38252443. DOI: 10.1001/jamaneurol.2023.5319.

[20] Gonzalez-Ortiz F, et al. Plasma phospho-tau in Alzheimer’s disease: towards diagnostic and therapeutic trial applications. Mol Neurodegener. 2023. PMID: 36927491. DOI: 10.1186/s13024-023-00605-8.

[21] Wang J, et al. Diagnostic accuracy of plasma p-tau217/Abeta42 for Alzheimer’s disease in clinical and community cohorts. Alzheimers Dement. 2025. PMID: 40156286.

[22] Verberk IMW, et al. Evidence-based standardized sample handling protocol for accurate blood-based Alzheimer’s disease biomarker measurement: results and consensus of the Global Biomarker Standardization Consortium. Alzheimers Dement. 2025. PMID: 41025225.

[23] De Meyer S, et al. Longitudinal associations of serum biomarkers with early cognitive, amyloid and grey matter changes. Brain. 2024. PMID: 37787146.

[24] Kjaergaard D, et al. Ethnic and racial influences on blood biomarkers for Alzheimer’s disease: a systematic review. J Alzheimers Dis. 2025. PMID: 39814528.

[25] Coleman A, et al. Evaluating finger-prick blood collection for remote quantification of neurofilament light in neurological diseases. J Neurol. 2025. PMID: 40637865.

[26] Geyer PE, et al. Plasma proteome profiling to detect and avoid sample-related biases in biomarker studies. EMBO Mol Med. 2019;11(11):e10427. PMID: 31566909. DOI: 10.15252/emmm.201910427.

[27] McKhann GM, et al. The diagnosis of dementia due to Alzheimer’s disease: recommendations from the National Institute on Aging-Alzheimer’s Association workgroups. Alzheimers Dement. 2011;7(3):263–269. PMID: 21514250. DOI: 10.1016/j.jalz.2011.03.005.

[28] Montalban X, et al. Diagnosis of multiple sclerosis: 2024 revisions of the McDonald criteria. Lancet Neurol. 2025;24(10):850–865. PMID: 40975101. DOI: 10.1016/S1474-4422(25)00270-4.

[29] Brooks BR, et al. El Escorial revisited: revised criteria for the diagnosis of amyotrophic lateral sclerosis. Amyotroph Lateral Scler Other Motor Neuron Disord. 2000;1(5):293–299. PMID: 11464847.

[30] Shefner JM, et al. A proposal for new diagnostic criteria for ALS. Clin Neurophysiol. 2020;131(8):1975–1978. PMID: 32387049. DOI: 10.1016/j.clinph.2020.04.005.

[31] Rascovsky K, et al. Sensitivity of revised diagnostic criteria for the behavioural variant of frontotemporal dementia. Brain. 2011;134(Pt 9):2456–2477. PMID: 21810890. DOI: 10.1093/brain/awr179.

[32] Yamada M, et al. Diagnostic Criteria for Dementia with Lewy Bodies: Updates and Future Directions. J Mov Disord. 2020;13(1):1–10. PMID: 31694357. DOI: 10.14802/jmd.19052.

[33] Pilotto A, Quaresima V, Trasciatti C, et al. Plasma p-tau217 in Alzheimer’s disease: Lumipulse and ALZpath SIMOA head-to-head comparison. Brain. 2025;148(2):408–415. DOI: 10.1093/brain/awae368.

[34] Martinuzzo C, et al. Multiplex plasma profiling of synaptic biomarkers in Alzheimer’s disease using NULISA: early alterations, APOE genotype effects, and pTau217 associations. medRxiv. 2026 [preprint]. DOI: 10.64898/2026.05.21.26353560.

