## Supplementary Table S1 for "Matrix matters: head-to-head concordance of serum and plasma for NULISAseq CNS Disease Panel"

**Supplementary Table S1. Per-protein serum-plasma concordance, paired test statistics and normality checks (130 proteins, non-bridged primary analysis)**

| Protein | n | t | df | p (t-test) | p_adj | Shapiro W | Shapiro p | Wilcoxon p | Spearman rho | Lin CCC | ICC(2,1) | dNPQ | CCC class |
| --- | --- | --- | --- | --- | --- | --- | --- | --- | --- | --- | --- | --- | --- |
| GDNF | 62 | -2.786 | 61 | 0.007 | 0.013 | 0.990 | 0.887 | 0.006 | 0.988 | 0.99 | 0.99 | -0.112 | Excellent |
| NEFH | 62 | -3.116 | 61 | 0.003 | 0.005 | 0.957 | 0.029 | 0.005 | 0.988 | 0.988 | 0.988 | -0.164 | Excellent |
| BASP1 | 62 | -2.764 | 61 | 0.008 | 0.013 | 0.934 | 0.002 | 0.014 | 0.972 | 0.986 | 0.986 | -0.108 | Excellent |
| IL17A | 62 | -3 | 61 | 0.004 | 0.007 | 0.972 | 0.159 | 0.007 | 0.961 | 0.977 | 0.977 | -0.111 | Excellent |
| VGF | 62 | -0.838 | 61 | 0.405 | 0.442 | 0.981 | 0.446 | 0.271 | 0.953 | 0.98 | 0.981 | -0.043 | Excellent |
| MSLN | 62 | -1.603 | 61 | 0.114 | 0.154 | 0.975 | 0.225 | 0.207 | 0.951 | 0.951 | 0.952 | -0.072 | Excellent |
| GFAP | 62 | 0.203 | 61 | 0.840 | 0.860 | 0.992 | 0.969 | 0.833 | 0.949 | 0.939 | 0.94 | 0.01 | Excellent |
| PDLIM5 | 62 | -0.112 | 61 | 0.912 | 0.919 | 0.925 | 9.88e-04 | 0.589 | 0.949 | 0.982 | 0.982 | -0.006 | Excellent |
| NEFL | 62 | -1.351 | 61 | 0.182 | 0.236 | 0.977 | 0.289 | 0.212 | 0.947 | 0.96 | 0.96 | -0.051 | Excellent |
| GDF15 | 62 | 8.796 | 61 | 1.90e-12 | 8.25e-12 | 0.768 | 1.62e-08 | 6.70e-11 | 0.941 | 0.918 | 0.919 | 0.535 | Excellent |
| UBB | 62 | -1.716 | 61 | 0.091 | 0.129 | 0.988 | 0.803 | 0.080 | 0.941 | 0.99 | 0.99 | -0.062 | Excellent |
| CRP | 62 | -1.121 | 61 | 0.266 | 0.330 | 0.936 | 0.003 | 0.199 | 0.933 | 0.958 | 0.959 | -0.054 | Excellent |
| IL5 | 62 | -0.916 | 61 | 0.363 | 0.412 | 0.883 | 2.61e-05 | 0.215 | 0.932 | 0.931 | 0.932 | -0.062 | Excellent |
| MME | 62 | -1.876 | 61 | 0.065 | 0.096 | 0.975 | 0.224 | 0.147 | 0.929 | 0.948 | 0.949 | -0.143 | Excellent |
| IL13 | 62 | -3.31 | 61 | 0.002 | 0.003 | 0.974 | 0.211 | 0.001 | 0.921 | 0.942 | 0.942 | -0.134 | Excellent |
| IFNG | 62 | -1.677 | 61 | 0.099 | 0.138 | 0.982 | 0.497 | 0.108 | 0.916 | 0.95 | 0.951 | -0.075 | Excellent |
| CRH | 62 | -9.744 | 61 | 4.76e-14 | 2.47e-13 | 0.947 | 0.009 | 5.24e-10 | 0.906 | 0.862 | 0.864 | -0.461 | Good |
| SAA1 | 62 | 8.448 | 61 | 7.49e-12 | 3.14e-11 | 0.965 | 0.070 | 5.00e-09 | 0.903 | 0.87 | 0.872 | 0.905 | Good |
| IL12p70 | 62 | -1.766 | 61 | 0.082 | 0.118 | 0.971 | 0.158 | 0.107 | 0.895 | 0.917 | 0.918 | -0.079 | Excellent |
| TREM2 | 62 | -0.705 | 61 | 0.483 | 0.519 | 0.989 | 0.846 | 0.556 | 0.889 | 0.916 | 0.917 | -0.029 | Excellent |
| pTau-231 | 62 | 13.45 | 61 | 6.61e-20 | 1.15e-18 | 0.952 | 0.016 | 7.77e-12 | 0.885 | 0.679 | 0.682 | 0.886 | Moderate |
| CHIT1 | 62 | -4.81 | 61 | 1.03e-05 | 2.79e-05 | 0.961 | 0.045 | 1.14e-05 | 0.883 | 0.965 | 0.966 | -0.325 | Excellent |
| CHI3L1 | 62 | -4.769 | 61 | 1.19e-05 | 3.16e-05 | 0.988 | 0.799 | 1.96e-05 | 0.877 | 0.825 | 0.827 | -0.294 | Good |
| pTau-181 | 62 | 10.922 | 61 | 5.48e-16 | 3.57e-15 | 0.965 | 0.078 | 3.30e-11 | 0.869 | 0.717 | 0.72 | 0.668 | Moderate |
| DDC | 62 | -7.553 | 61 | 2.59e-10 | 9.35e-10 | 0.986 | 0.676 | 2.30e-08 | 0.861 | 0.827 | 0.829 | -0.278 | Good |
| pTau-217 | 62 | 11.094 | 61 | 2.89e-16 | 2.35e-15 | 0.904 | 1.47e-04 | 2.48e-11 | 0.846 | 0.699 | 0.703 | 0.644 | Moderate |
| CXCL10 | 62 | 1.073 | 61 | 0.288 | 0.353 | 0.939 | 0.004 | 0.145 | 0.844 | 0.892 | 0.894 | 0.059 | Good |
| PDGFRB | 62 | -2.091 | 61 | 0.041 | 0.063 | 0.987 | 0.732 | 0.039 | 0.842 | 0.874 | 0.876 | -0.101 | Good |
| FABP3 | 62 | -2.12 | 61 | 0.038 | 0.060 | 0.972 | 0.172 | 0.054 | 0.833 | 0.872 | 0.874 | -0.103 | Good |
| BD-pTau-181 | 62 | 13.429 | 61 | 7.10e-20 | 1.15e-18 | 0.975 | 0.229 | 8.99e-12 | 0.82 | 0.641 | 0.644 | 0.962 | Moderate |
| IL2 | 62 | -1.057 | 61 | 0.295 | 0.358 | 0.968 | 0.100 | 0.293 | 0.817 | 0.85 | 0.852 | -0.054 | Good |
| KDR | 62 | -1.558 | 61 | 0.124 | 0.167 | 0.981 | 0.471 | 0.097 | 0.804 | 0.972 | 0.972 | -0.095 | Excellent |
| POSTN | 62 | 2.031 | 61 | 0.047 | 0.070 | 0.968 | 0.108 | 0.020 | 0.794 | 0.803 | 0.806 | 0.14 | Good |
| CCL11 | 62 | -1.542 | 61 | 0.128 | 0.170 | 0.973 | 0.182 | 0.197 | 0.777 | 0.8 | 0.802 | -0.089 | Good |
| IL18 | 62 | 0.456 | 61 | 0.650 | 0.682 | 0.693 | 4.25e-10 | 0.377 | 0.777 | 0.546 | 0.55 | 0.041 | Moderate |
| KLK6 | 62 | -1.62 | 61 | 0.110 | 0.151 | 0.953 | 0.019 | 0.347 | 0.777 | 0.772 | 0.775 | -0.074 | Good |
| IL10 | 62 | -2.725 | 61 | 0.008 | 0.015 | 0.924 | 9.13e-04 | 0.010 | 0.773 | 0.802 | 0.805 | -0.176 | Good |
| IL9 | 62 | -12.653 | 61 | 1.04e-18 | 1.13e-17 | 0.964 | 0.065 | 1.77e-11 | 0.771 | 0.562 | 0.566 | -0.859 | Moderate |
| BD-pTau-217 | 62 | 7.253 | 61 | 8.50e-10 | 2.99e-09 | 0.892 | 5.15e-05 | 7.15e-10 | 0.77 | 0.76 | 0.763 | 0.301 | Good |
| FOLR1 | 62 | -1.017 | 61 | 0.313 | 0.367 | 0.975 | 0.244 | 0.197 | 0.766 | 0.746 | 0.749 | -0.042 | Moderate |
| SFTPD | 62 | -5.07 | 61 | 3.96e-06 | 1.12e-05 | 0.990 | 0.913 | 1.10e-05 | 0.761 | 0.712 | 0.715 | -0.344 | Moderate |
| SMOC1 | 62 | -5.507 | 61 | 7.72e-07 | 2.28e-06 | 0.986 | 0.696 | 1.27e-06 | 0.76 | 0.652 | 0.656 | -0.413 | Moderate |
| IL6 | 62 | -2.865 | 61 | 0.006 | 0.010 | 0.667 | 1.40e-10 | 0.002 | 0.747 | 0.651 | 0.655 | -0.252 | Moderate |
| GDI1 | 62 | 1.199 | 61 | 0.235 | 0.294 | 0.990 | 0.910 | 0.283 | 0.73 | 0.934 | 0.935 | 0.066 | Excellent |
| VSNL1 | 62 | 2.054 | 61 | 0.044 | 0.068 | 0.984 | 0.579 | 0.016 | 0.73 | 0.792 | 0.794 | 0.074 | Good |
| APOE | 62 | -7.821 | 61 | 8.95e-11 | 3.42e-10 | 0.974 | 0.220 | 4.79e-09 | 0.722 | 0.626 | 0.63 | -0.329 | Moderate |
| ICAM1 | 62 | -1.81 | 61 | 0.075 | 0.109 | 0.963 | 0.060 | 0.102 | 0.715 | 0.707 | 0.711 | -0.091 | Moderate |
| NPTXR | 62 | -1.654 | 61 | 0.103 | 0.143 | 0.957 | 0.029 | 0.095 | 0.715 | 0.685 | 0.688 | -0.089 | Moderate |
| CCL22 | 62 | -3.376 | 61 | 0.001 | 0.003 | 0.950 | 0.013 | 0.002 | 0.711 | 0.727 | 0.73 | -0.187 | Moderate |
| CNTN2 | 62 | -2.153 | 61 | 0.035 | 0.056 | 0.989 | 0.861 | 0.038 | 0.71 | 0.72 | 0.724 | -0.161 | Moderate |
| NGF | 62 | 3.676 | 61 | 5.02e-04 | 0.001 | 0.980 | 0.397 | 2.46e-04 | 0.693 | 0.723 | 0.726 | 0.118 | Moderate |
| IL4 | 62 | -10.545 | 61 | 2.25e-15 | 1.27e-14 | 0.977 | 0.287 | 2.67e-10 | 0.689 | 0.401 | 0.405 | -1.334 | Poor |
| VCAM1 | 62 | -1.201 | 61 | 0.234 | 0.294 | 0.990 | 0.904 | 0.236 | 0.682 | 0.702 | 0.705 | -0.04 | Moderate |
| IGF1R | 62 | -4.042 | 61 | 1.51e-04 | 3.57e-04 | 0.969 | 0.116 | 8.68e-04 | 0.677 | 0.557 | 0.561 | -0.135 | Moderate |
| CSF2 | 62 | -2.586 | 61 | 0.012 | 0.020 | 0.773 | 2.07e-08 | 0.005 | 0.675 | 0.641 | 0.645 | -0.17 | Moderate |
| CCL4 | 62 | -10.947 | 61 | 4.99e-16 | 3.42e-15 | 0.906 | 1.77e-04 | 1.69e-11 | 0.673 | 0.509 | 0.513 | -1.215 | Moderate |
| FCN2 | 62 | -0.962 | 61 | 0.340 | 0.391 | 0.860 | 4.43e-06 | 0.344 | 0.671 | 0.53 | 0.534 | -0.099 | Moderate |
| BD-pTau-231 | 62 | 7.57 | 61 | 2.42e-10 | 9.00e-10 | 0.972 | 0.158 | 6.43e-09 | 0.666 | 0.593 | 0.597 | 0.203 | Moderate |
| CX3CL1 | 62 | -0.844 | 61 | 0.402 | 0.442 | 0.972 | 0.161 | 0.207 | 0.664 | 0.688 | 0.691 | -0.045 | Moderate |
| BACE1 | 62 | -2.652 | 61 | 0.010 | 0.017 | 0.988 | 0.800 | 0.011 | 0.662 | 0.64 | 0.644 | -0.09 | Moderate |
| REST | 62 | 0.408 | 61 | 0.685 | 0.712 | 0.796 | 7.69e-08 | 0.624 | 0.661 | 0.488 | 0.492 | 0.032 | Poor |
| NPTX1 | 62 | -2.186 | 61 | 0.033 | 0.052 | 0.979 | 0.350 | 0.046 | 0.66 | 0.637 | 0.641 | -0.115 | Moderate |
| ACHE | 62 | -2.294 | 61 | 0.025 | 0.041 | 0.976 | 0.275 | 0.030 | 0.659 | 0.693 | 0.697 | -0.107 | Moderate |
| IL33 | 62 | 4.881 | 61 | 7.95e-06 | 2.20e-05 | 0.844 | 1.53e-06 | 9.17e-08 | 0.655 | 0.475 | 0.48 | 0.433 | Poor |
| IGFBP7 | 62 | -0.908 | 61 | 0.367 | 0.412 | 0.964 | 0.068 | 0.721 | 0.652 | 0.564 | 0.568 | -0.039 | Moderate |
| TREM1 | 62 | -8.248 | 61 | 1.65e-11 | 6.49e-11 | 0.982 | 0.489 | 5.91e-09 | 0.649 | 0.529 | 0.533 | -0.47 | Moderate |
| IL6R | 62 | -3.259 | 61 | 0.002 | 0.004 | 0.959 | 0.038 | 0.007 | 0.648 | 0.613 | 0.617 | -0.158 | Moderate |
| NPTX2 | 62 | -2.441 | 61 | 0.018 | 0.029 | 0.995 | 0.997 | 0.020 | 0.642 | 0.651 | 0.654 | -0.124 | Moderate |
| HBA1 | 62 | 0.963 | 61 | 0.339 | 0.391 | 0.935 | 0.003 | 0.060 | 0.632 | 0.525 | 0.529 | 0.232 | Moderate |
| CST3 | 62 | 1.016 | 61 | 0.314 | 0.367 | 0.969 | 0.124 | 0.207 | 0.626 | 0.712 | 0.715 | 0.026 | Moderate |
| IL16 | 62 | -3.788 | 61 | 3.49e-04 | 7.83e-04 | 0.896 | 7.16e-05 | 1.52e-05 | 0.626 | 0.482 | 0.486 | -0.342 | Poor |
| SLIT2 | 62 | -3.885 | 61 | 2.55e-04 | 5.92e-04 | 0.972 | 0.175 | 2.32e-04 | 0.625 | 0.532 | 0.536 | -0.175 | Moderate |
| PGF | 62 | -3.427 | 61 | 0.001 | 0.002 | 0.983 | 0.532 | 0.003 | 0.615 | 0.592 | 0.596 | -0.146 | Moderate |
| CALB2 | 62 | -0.578 | 61 | 0.565 | 0.598 | 0.972 | 0.167 | 0.533 | 0.611 | 0.688 | 0.692 | -0.039 | Moderate |
| TAFA5 | 62 | 9.695 | 61 | 5.75e-14 | 2.88e-13 | 0.980 | 0.401 | 4.58e-10 | 0.589 | 0.404 | 0.408 | 1.121 | Poor |
| MAPT | 62 | 12.802 | 61 | 6.19e-19 | 8.05e-18 | 0.943 | 0.006 | 9.91e-12 | 0.587 | 0.289 | 0.293 | 1.098 | Poor |
| IL15 | 62 | -3.09 | 61 | 0.003 | 0.006 | 0.972 | 0.169 | 8.90e-04 | 0.576 | 0.522 | 0.526 | -0.124 | Moderate |
| S100B | 62 | -3.211 | 61 | 0.002 | 0.004 | 0.731 | 2.41e-09 | 4.10e-06 | 0.556 | 0.438 | 0.442 | -0.303 | Poor |
| AGRN | 62 | -1.979 | 61 | 0.052 | 0.078 | 0.990 | 0.882 | 0.049 | 0.549 | 0.515 | 0.519 | -0.094 | Moderate |
| CD63 | 62 | -11.886 | 61 | 1.60e-17 | 1.39e-16 | 0.983 | 0.558 | 6.39e-11 | 0.539 | 0.277 | 0.281 | -0.712 | Poor |
| BD-MAPT | 62 | 7.143 | 61 | 1.32e-09 | 4.50e-09 | 0.969 | 0.123 | 7.29e-09 | 0.532 | 0.545 | 0.549 | 0.152 | Moderate |
| TEK | 62 | -2.876 | 61 | 0.006 | 0.010 | 0.961 | 0.048 | 0.003 | 0.522 | 0.455 | 0.459 | -0.14 | Poor |
| GOT1 | 62 | -1.883 | 61 | 0.065 | 0.095 | 0.983 | 0.538 | 0.074 | 0.515 | 0.501 | 0.505 | -0.116 | Moderate |
| CCL26 | 62 | -8.29 | 61 | 1.40e-11 | 5.67e-11 | 0.987 | 0.745 | 3.41e-09 | 0.498 | 0.365 | 0.369 | -0.817 | Poor |
| CCL13 | 62 | -11.036 | 61 | 3.59e-16 | 2.75e-15 | 0.978 | 0.338 | 9.72e-11 | 0.491 | 0.264 | 0.267 | -1.094 | Poor |
| UCHL1 | 62 | 1.024 | 61 | 0.310 | 0.367 | 0.984 | 0.588 | 0.319 | 0.491 | 0.537 | 0.541 | 0.016 | Moderate |
| CCL17 | 62 | -13.47 | 61 | 6.16e-20 | 1.15e-18 | 0.983 | 0.548 | 1.46e-11 | 0.47 | 0.171 | 0.173 | -2.645 | Poor |
| PTN | 62 | -1.047 | 61 | 0.299 | 0.360 | 0.963 | 0.056 | 0.392 | 0.468 | 0.969 | 0.969 | -0.023 | Excellent |
| VEGFD | 62 | -10.671 | 61 | 1.40e-15 | 8.30e-15 | 0.975 | 0.228 | 1.02e-10 | 0.458 | 0.203 | 0.206 | -1.313 | Poor |
| PSEN1 | 62 | -4.367 | 61 | 4.97e-05 | 1.24e-04 | 0.876 | 1.48e-05 | 5.01e-06 | 0.455 | 0.252 | 0.255 | -0.491 | Poor |
| TNF | 62 | -6.182 | 61 | 5.78e-08 | 1.88e-07 | 0.827 | 5.02e-07 | 5.67e-09 | 0.455 | 0.233 | 0.236 | -0.571 | Poor |
| IL1B | 62 | -2.494 | 61 | 0.015 | 0.026 | 0.921 | 6.48e-04 | 0.003 | 0.454 | 0.632 | 0.636 | -0.318 | Moderate |
| Aβ42 | 62 | 9.619 | 61 | 7.71e-14 | 3.71e-13 | 0.918 | 5.11e-04 | 1.02e-09 | 0.452 | 0.236 | 0.239 | 0.887 | Poor |
| SQSTM1 | 62 | -3.592 | 61 | 6.55e-04 | 0.001 | 0.948 | 0.011 | 0.008 | 0.441 | 0.453 | 0.457 | -0.324 | Poor |
| ARSA | 62 | 4.655 | 61 | 1.80e-05 | 4.67e-05 | 0.973 | 0.194 | 9.42e-05 | 0.436 | 0.298 | 0.301 | 0.994 | Poor |
| SNCB | 62 | 3.36 | 61 | 0.001 | 0.003 | 0.974 | 0.220 | 0.001 | 0.425 | 0.372 | 0.376 | 0.082 | Poor |
| FGF2 | 62 | -3.437 | 61 | 0.001 | 0.002 | 0.968 | 0.104 | 5.20e-04 | 0.418 | 0.341 | 0.345 | -0.717 | Poor |
| TIMP3 | 62 | -12.158 | 61 | 6.02e-18 | 6.02e-17 | 0.986 | 0.701 | 5.55e-11 | 0.417 | 0.154 | 0.156 | -2.701 | Poor |
| CXCL1 | 62 | -12.856 | 61 | 5.11e-19 | 7.39e-18 | 0.962 | 0.050 | 7.02e-11 | 0.416 | 0.217 | 0.22 | -1.774 | Poor |
| CCL2 | 62 | -10.746 | 61 | 1.06e-15 | 6.54e-15 | 0.827 | 4.75e-07 | 1.33e-11 | 0.414 | 0.099 | 0.101 | -1.338 | Poor |
| NPY | 62 | 11 | 61 | 4.10e-16 | 2.96e-15 | 0.926 | 0.001 | 1.02e-10 | 0.395 | 0.131 | 0.133 | 2.868 | Poor |
| PGK1 | 62 | -4.44 | 61 | 3.85e-05 | 9.80e-05 | 0.941 | 0.005 | 1.34e-05 | 0.378 | 0.333 | 0.336 | -1.002 | Poor |
| MDH1 | 62 | -0.035 | 61 | 0.972 | 0.972 | 0.983 | 0.526 | 0.905 | 0.363 | 0.4 | 0.404 | -0.006 | Poor |
| YWHAZ | 62 | -0.71 | 61 | 0.481 | 0.519 | 0.847 | 1.82e-06 | 0.101 | 0.357 | 0.556 | 0.56 | -0.05 | Moderate |
| S100A12 | 62 | -9.188 | 61 | 4.11e-13 | 1.91e-12 | 0.903 | 1.31e-04 | 6.70e-09 | 0.354 | 0.191 | 0.193 | -1.838 | Poor |
| PRDX6 | 62 | -1.23 | 61 | 0.223 | 0.287 | 0.984 | 0.601 | 0.274 | 0.346 | 0.345 | 0.349 | -0.218 | Poor |
| RUVBL2 | 62 | -10.36 | 61 | 4.53e-15 | 2.45e-14 | 0.961 | 0.044 | 2.33e-10 | 0.333 | 0.094 | 0.095 | -0.918 | Poor |
| FLT1 | 62 | -0.908 | 61 | 0.368 | 0.412 | 0.992 | 0.967 | 0.347 | 0.32 | 0.33 | 0.333 | -0.063 | Poor |
| CCL3 | 62 | -6.786 | 61 | 5.41e-09 | 1.80e-08 | 0.796 | 7.47e-08 | 2.22e-10 | 0.318 | 0.127 | 0.129 | -1.094 | Poor |
| TARDBP | 62 | -3.853 | 61 | 2.83e-04 | 6.46e-04 | 0.965 | 0.074 | 1.09e-04 | 0.318 | 0.209 | 0.212 | -0.629 | Poor |
| ANXA5 | 62 | 13.938 | 61 | 1.27e-20 | 3.30e-19 | 0.862 | 5.27e-06 | 7.77e-12 | 0.273 | 0.08 | 0.081 | 4.486 | Poor |
| YWHAG | 62 | -0.132 | 61 | 0.896 | 0.910 | 0.990 | 0.889 | 0.894 | 0.272 | 0.301 | 0.304 | -0.005 | Poor |
| HTT | 62 | -0.849 | 61 | 0.399 | 0.442 | 0.979 | 0.364 | 0.225 | 0.269 | 0.247 | 0.25 | -0.153 | Poor |
| Aβ40 | 62 | 5.193 | 61 | 2.52e-06 | 7.27e-06 | 0.900 | 1.05e-04 | 5.77e-07 | 0.26 | 0.163 | 0.165 | 1.027 | Poor |
| Oligo-SNCA | 62 | -0.338 | 61 | 0.736 | 0.760 | 0.990 | 0.881 | 0.774 | 0.248 | 0.296 | 0.299 | -0.12 | Poor |
| CXCL8 | 62 | -8.909 | 61 | 1.22e-12 | 5.48e-12 | 0.904 | 1.47e-04 | 1.41e-10 | 0.234 | 0.241 | 0.244 | -2.063 | Poor |
| IL7 | 62 | -15.356 | 61 | 1.25e-22 | 5.43e-21 | 0.949 | 0.012 | 3.00e-11 | 0.232 | 0.031 | 0.032 | -2.412 | Poor |
| SNCA | 62 | -0.605 | 61 | 0.547 | 0.583 | 0.987 | 0.732 | 0.561 | 0.226 | 0.248 | 0.251 | -0.096 | Poor |
| BDNF | 62 | -12.744 | 61 | 7.58e-19 | 8.95e-18 | 0.989 | 0.832 | 3.30e-11 | 0.223 | 0.027 | 0.027 | -2.693 | Poor |
| SNAP25 | 62 | 5.868 | 61 | 1.94e-07 | 6.02e-07 | 0.903 | 1.29e-04 | 1.51e-07 | 0.219 | 0.078 | 0.079 | 0.077 | Poor |
| ENO2 | 62 | 24.212 | 61 | 5.54e-33 | 7.20e-31 | 0.940 | 0.005 | 8.56e-12 | 0.204 | 0.019 | 0.019 | 5.716 | Poor |
| pTDP43-409 | 62 | 1.38 | 61 | 0.173 | 0.227 | 0.939 | 0.004 | 0.250 | 0.192 | 0.193 | 0.196 | 0.06 | Poor |
| VEGFA | 62 | -12.129 | 61 | 6.67e-18 | 6.20e-17 | 0.991 | 0.918 | 2.15e-11 | 0.168 | 0.048 | 0.049 | -0.916 | Poor |
| NRGN | 62 | 14.01 | 61 | 9.99e-21 | 3.25e-19 | 0.984 | 0.602 | 1.86e-11 | 0.147 | 0.052 | 0.053 | 4.391 | Poor |
| pSNCA-129 | 62 | -6.078 | 61 | 8.63e-08 | 2.74e-07 | 0.975 | 0.228 | 6.20e-07 | 0.14 | 0.09 | 0.091 | -1.16 | Poor |
| PARK7 | 62 | -4.094 | 61 | 1.27e-04 | 3.07e-04 | 0.977 | 0.305 | 1.37e-04 | 0.122 | 0.205 | 0.207 | -0.715 | Poor |
| CD40LG | 62 | -19.606 | 61 | 5.06e-28 | 3.29e-26 | 0.927 | 0.001 | 9.91e-12 | 0.102 | 0.003 | 0.003 | -4.743 | Poor |
| SOD1 | 62 | -4.092 | 61 | 1.28e-04 | 3.07e-04 | 0.991 | 0.916 | 1.97e-04 | 0.073 | 0.087 | 0.089 | -0.788 | Poor |
| SFRP1 | 62 | 5.698 | 61 | 3.74e-07 | 1.13e-06 | 0.973 | 0.187 | 3.96e-06 | 0.07 | 0.039 | 0.039 | 1.473 | Poor |
| Aβ38 | 62 | 1.224 | 61 | 0.226 | 0.287 | 0.939 | 0.004 | 0.523 | 0.01 | -0.03 | -0.031 | 0.294 | Poor |

**Legend.** n = valid paired samples (both matrices above LOD); t, df = two-tailed paired t-test on log2 NPQ (df = n-1); p (t-test) = exact paired t-test p; p_adj = Benjamini-Hochberg FDR; Shapiro W / Shapiro p = Shapiro-Wilk test of normality of the paired differences; Wilcoxon p = exact Wilcoxon signed-rank p (distribution-free sensitivity analysis); Spearman rho = rank concordance; Lin CCC = concordance correlation coefficient; ICC(2,1) = two-way random-effects absolute-agreement intraclass correlation; dNPQ = mean paired plasma - serum difference (NPQ units). CCC class: Excellent >=0.90; Good 0.80-<0.90; Moderate 0.65-<0.80; Poor <0.65. Ordered by descending Spearman rho.
